# Understanding the impact of the COVID-19 pandemic on a socially deprived UK coastal town: a preliminary exploratory analysis of health and socioeconomic data

**DOI:** 10.1101/2021.12.22.21268232

**Authors:** Maddy French, Mark Spencer, Mike Walker, Afzal Patel, Neil Clarke, Ross Hughes, Collette Taylor, Margaret Orwin, Alicia Elliott, Karl Worsley, Julie Casson, Stephen Milan, Mark Bowen, Luigi Sedda

## Abstract

**Introduction:** In addition to the direct impact of COVID-19 infections on health and mortality, a growing body of literature indicates there are wide-ranging indirect impacts of the COVID-19 pandemic and associated public health measures on population health and wellbeing. Exploring these indirect impacts in the context of a socially deprived UK coastal town will help identify priority areas to focus COVID-19 recovery efforts on.

**Methods:** Data on primary care diagnosis, hospital admissions, and several socioeconomic outcomes between 2016 and Spring 2021 in the UK town of Fleetwood were collected and analysed in an exploratory analysis looking at pre- and post-COVID-19 patterns in health and social outcomes. Weekly and monthly trends were plotted by time and differences between periods examined using Chi-squared and t-tests.

**Results:** Initial falls in hospital admissions and diagnoses of conditions in primary care in March 2020 were followed by sustained changes to health service activity for specific diagnostic and demographic groups, including for chronic kidney disease and young people. Increases in the number of people receiving Universal Credit and children eligible for free school meals appear to be greater for those in the least deprived areas of the town.

**Discussion:** These exploratory findings provide initial evidence of the sustained impact of the pandemic across several health and social outcomes. Examining these trends in multivariate analyses will further test these associations and establish the strength of the medium term impact of the pandemic on the population of this coastal town. Advanced modelling of this data is ongoing and will be published shortly.

## Introduction

The arrival of COVID-19 has exposed and likely exacerbated decades of worsening health inequalities in many UK towns (1–3). For communities already tackling persistent and entrenched health and socioeconomic inequities, the pandemic has created additional challenges but may also offer opportunities for reform and change. As well as COVID-19 infections having a direct impact on health and mortality, a growing body of literature suggests there are wide-ranging indirect impacts of the pandemic and associated public health measures on health and wellbeing (4–6).

The arrival of COVID-19 necessitated a substantial shift in focus within healthcare towards the care of people with acute COVID-19, and the protection of other patients and staff from infection. As would be expected, this had an indirect yet substantial impact on the patterns of healthcare use for other patients. Someof the reported short-term effects of the pandemic include an immediate drop in use of hospital and primary care services as people avoided non-urgent healthcare, and some healthcare services were paused, during the early months of the pandemic (4,5,7). The pause in access may also have indirect longer-te rm impacts on people with chronic health conditions. A small survey of Chronic Obstructive Pulmonary Disease (COPD) patients, for example, found that many experienced worsening of their COPD control during the ‘lockdown’ period (8). Other chronic conditions such as diabetes may have worsened for some people through the negative impact of reduced physical inactivity and increased food intake (9).

Several studies suggest the COVID-19 pandemic also had an immediate impact on healthcare provided to people with mental health needs in the UK. Primary care contacts for people with acute mental health conditions in the UK dropped considerably during this time, and for most conditions had not recovered to normal levels by July 2020 (4). The incidence of depression, anxiety disorders and first antidepressant prescribing was lower in April 2020 compared to previous years; by September these had returned to approximately normal levels in England but remained low in the rest of the UK (12).

Alongside these patterns in appearances at health services, the mental health needs of some population groups may have increased. The combination of the social determinants of mental health with pandemic-related stressors is likely to exacerbate or create mental health burdens for marginalised, discriminated, or socially disadvantaged populations (13). There is consistent evidence suggesting the pandemic had a detrimental effect on mental health, particularly among women, young people, and those who were already financially insecure prior to the crisis (6,14,15).

At the same time COVID-19 impacted health and healthcare, it had an impact on the socioeconomic conditions of the communities affected by the pandemic. With a high share of employment sitting within the accommodation, leisure and tourism sectors, the UK is a country vulnerable to the negative economic impacts of the pandemic (16). While it is too early to estimate the long term impacts of the pandemic on household incomes, evidence from the 2008 global financial crash suggests that poverty began to increase several years after the crisis in reaction to austerity, highlightingthe importance of policy responses to crises (17). Following the arrival of COVID-19 in the UK, attempts were made to cushion the economic impacts of the public health measures through the Job Retention Scheme and an uplift to the Universal Credit payments for those in unemployment (18). Nonetheless, some population groups experienced, and likely continue to experience, greater impacts on their finances. Low income families with young children reported increases in household expenditure in 2020, and long-term forecasts suggest more parents, and thus children, will be pushed into precarious financial circumstances (19).

A potential consequence of these economic impacts includes increased food insecurity and poor health. A survey tracking food standards in the UK during Covid-19 suggests that in March 2021 the level of food insecurity remained high, with 17% of respondents skipping meals or cutting the size of meals because they did not have enough money to buy food (20). Well established links between unemployment and poor health have also led to predictions of increased physical and mental health burdens potentially arising from pandemic-related unemployment (21,22).

The aim of this analysis is to explore the indirect impacts the COVID-19 pandemic has had on health and socioeconomic outcomes in a socially deprived coastal town in North West England. In this study we provide a preliminary assessment of the post-COVID-19 emerging trends for health and socioeconomic conditions in Fleetwood. This preliminary assessment will be followed by a robust analysis of the spatial and temporal changes in outcomes between pre- and post-COVID-19 pandemic period, including the effect of different levels of deprivation in Fleetwood on these changes. Better understanding of the extent of the impact of the pandemic will help Fleetwood and similar coastal communities to prioritise recovery efforts for those most in need of support.

## Methods

### Study setting

Fleetwood is a coastal town in Lancashire home to approximately 30,000 people, many of whom were already experiencing poor health and living in disadvantaged socioeconomic circumstances prior to the pandemic (23). Forty-three percent of neighbourhoods in Fleetwood are amongst some of the 10% most deprived areas in the country and, like other deprived areas, experience some of the associated poor physical and mental health outcomes (24).

As a coastal community without a large employer or industry, Fleetwood is a place that is geographically and economically ‘on the periphery’, vulnerable to feeling the worst effects of phenomena such as financial crises and pandemics (25,26). It also has a population older than the English average, with 22% of its population aged over 65-years-old, compared to 18% nationally (27). A report released early on in the pandemic suggested that people in coastal areas were particularly vulnerable to the health and welfare effects of COVID-19 (2 8). However, coastal towns also offer opportunitiesto improve health and wellbeing, with good air quality and access to outdoors providing some protective effects against poor health (29).

### Data collection

Data were originally collected as part of routine clinical care or routine service delivery and surveillance in Fleetwood (Table 1). Information about diagnosis in primary care and hospital admissions for patients registered at a Fleetwood GP practice was extracted and provided to the research team by the Midlands and Lancashire Commissioning Support Unit. These datasets included information on patient age band, sex, the Lower Layer Super Output Area (LSOA) for the area where the patient lived, and date of activity. Data from primary care were provided for the period April 2016 to June 2021. The dataset for hospital admissions also included the admission outcome, diagnosis, and secondary diagnoses upon admission. Data on hospital admissions were provided for the period April 2016 to April 2021.

**Table 1:** Percentage of Fleetwood population by income deprivation area

| Income Decile | Number of LSOA areas | Income deprived | Percentage (%) |
| --- | --- | --- | --- |
| 1 | 43 | Most | 52 |
| 2 | 9 |  |  |
| 3 | 4 | Least | 49 |
| 5 | 19 |  |  |
| 6 | 14 |  |  |
| 7 | 11 |  |  |

Data on the number of young people on apprenticeships or classed as Not in Education, Employment, or Training (NEET), and the number of pupils eligible to receive free school meals, were provided by Lancashire County Council, aggregated by month and area. Information about street crime (number of crimes by month and LSOA) were extracted by the research team from open access datasets published by the police(www.data.police.uk) for the period June 2018 - June 2021 (historic data are only available up to two years). The number of unemployment claimants in Fleetwood aggregated by month, LSOA, age band, and sex were extracted from the open access dataset published by the Department for Work and Pensions (www.stat-xplore.dwp.gov.uk). Data on LSOA and Fleetwood population sizeby sex and age group were downloaded from open access datasets published by the Office for National Statistics (www.nomisweb.co.uk).

Information about area income deprivation was collected from the English indices of deprivation 2019 (24). The indices of deprivation include the Index of Multiple Deprivation, a composite measure of area deprivation, made up of individual domains of deprivation across income, employment, education, health, crime, environment, and access to services. In this study, data on employment, education, health, and crime in the local area was already collected, meaning it was appropriate to use only the income deprivation domain from the indices of deprivation. Using one domain also offers greater conceptual clarity than the composite Index of Multiple Deprivation, which includes data on multiple mechanisms between socioeconomic characteristics and health outcomes.

The distribution of area income deprivation in Fleetwood is shown in Table 1. In this study, small areas aggregate into two categories: the most income deprived areas (deciles 1 and 2) and the least income deprived areas (deciles 3,5-7). There are no areas in income decile 4 in Fleetwood. Income deprivation is subsequently used as a dichotomous variable.

### Statistical analysis

An exploratory analysis was conducted to look at the associations between health and socioeconomic outcomes, the pandemic, and selected patient or population demographics. To examine differences between the pre- and post-pandemic periods, a dummy variable for pre-March 2020 (0s) and post-March 2020 (1s) was created. Where data was provided by week, the week beginning 16th March 2020 was taken as the start of the pandemic in Fleetwood. For monthly aggregated data, the beginning of the pandemic was taken as 1st March 2020.

Weekly hospital admission rates stratified by age were created using Fleetwood population estimates for age bands 0-14, 15-24, 25-44, 45-64, 65-84, and 85+ as denominators. To calculate weekly rates of hospital admissions stratified by diagnosis, and of new diagnoses or consultations in primary care, the total Fleetwood population estimate was used. Total Fleetwood population estimates were available for years 2016-2019. Population rates for 2020 and 2021 were created using the 2019 population as the denominator.

For monthly rates of Universal Credit claimants in Fleetwood stratified by age, population estimates were calculated using population estimates for age bands 16-24, 25-54, and 55+ for years 2016-2021, which cover labour market age groups. Population rates for 2020 and 2021 were created using the 2019 population as the denominator. Young adults who can be classed as NEET must be aged between 16 and 24-year-olds. However, population estimates are only available for 15-17 and 18-24-year-olds. To create rates of young people on apprenticeships or NEET, population estimates of 15-24 year olds were used as a denominator

Rates and counts were plotted with time (month or week depending on data availability). Differences between the pre-March 2020 and post-March 2020 periods were tested using t-tests (continuous data) or Chi-squared tests (categorical data) when looking at group differences, with a p-value *<*0.05 threshold for statistical significance.

## Results

### Hospital admissions

The unadjusted trend in overall hospital admissions of residents in Fleetwood is shown in Figure 1. These plots suggest there was a fall in hospital admissions following the onset of the COVID-19 pandemic in March 2020 and low levels of overall hospital admissions have been sustained in the months since. The demographics patterns of patients being admitted to hospital changed following the pandemic for some patient groups. While the distribution of hospital admissions between patients of different sexes, or from difference income deprivation areas, remained similar in the post-March 2020 period (not shown), there were changes in the age demographics of patients being admitted to hospital (Figure 1). Figure 1 shows the time series trend in hospital admissions broken down by six age bands, indicating that admissions for patients aged between 0 and 24-years-old saw substantial falls.

**Figure 1:**
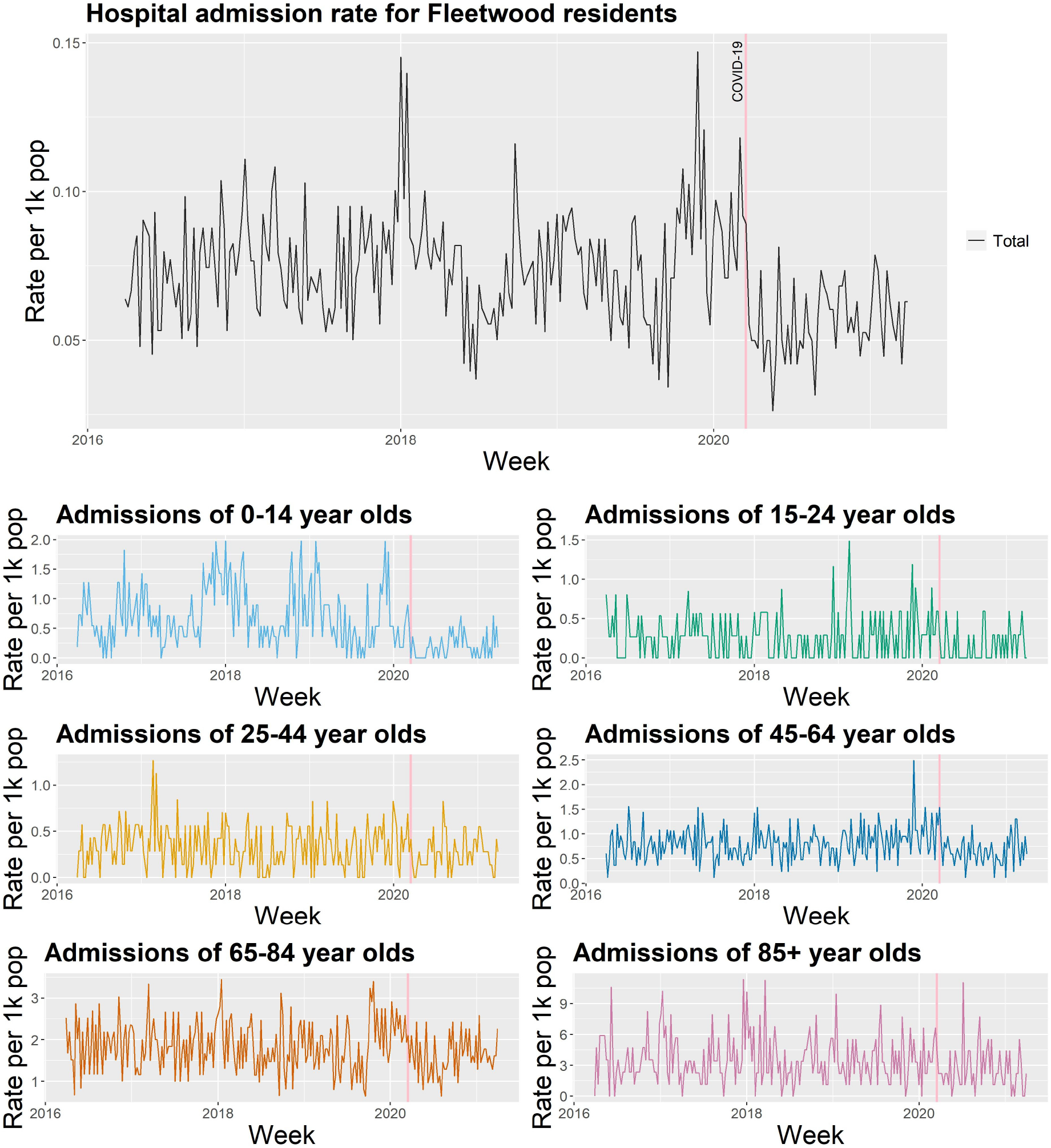
Weekly hospital admission rates stratified by age group

### Hospital admissions by diagnosis

Changes in hospital admission rates in the post-March 2020 period also differed depending on the primary diagnosis patients had upon admission. The time series trends of hospital admission rates for patients with a primary diagnosis of a respiratory condition, cardiovascular disease, and mental health are shown in Figure 2, as examples of some of the different trends in the post-March 2020 period.

**Figure 2:**
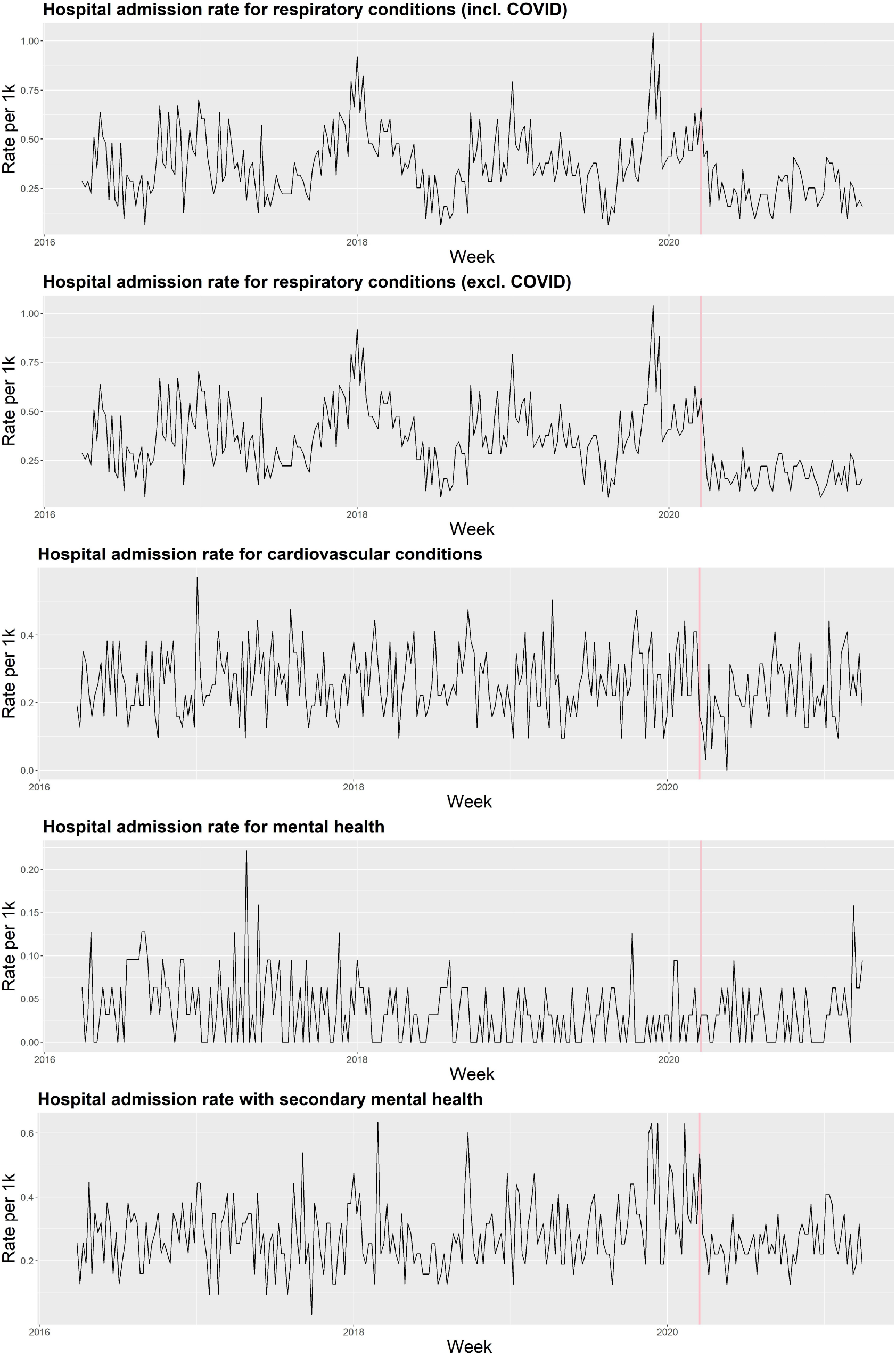
Weekly hospital admission rates stratified by selected primary diagnosis groups

#### Admissions with a primary respiratory diagnosis

Immediate falls in admissions where the primary diagnosis was respiratory-related in Spring 2020 were followed by sustained low admission rates up until the most recent week data were available for. As indicated in Figure 2 this remains the case even when COVID-19 admissions are included. Therefore it is unlikely that the fall in admissions for reasons relating to COPD and pneumonia was offset by the same patients being admitted with COVID-19, although there is likely to be some overlap. Prior to March 2020, admissions with a primary respiratory diagnosis accounted for 43% of all admissions, dropping in post-March 2020 to 37% (including COVID-19) and 27% (excluding COVID-19) (Table 3).

**Table 2:**
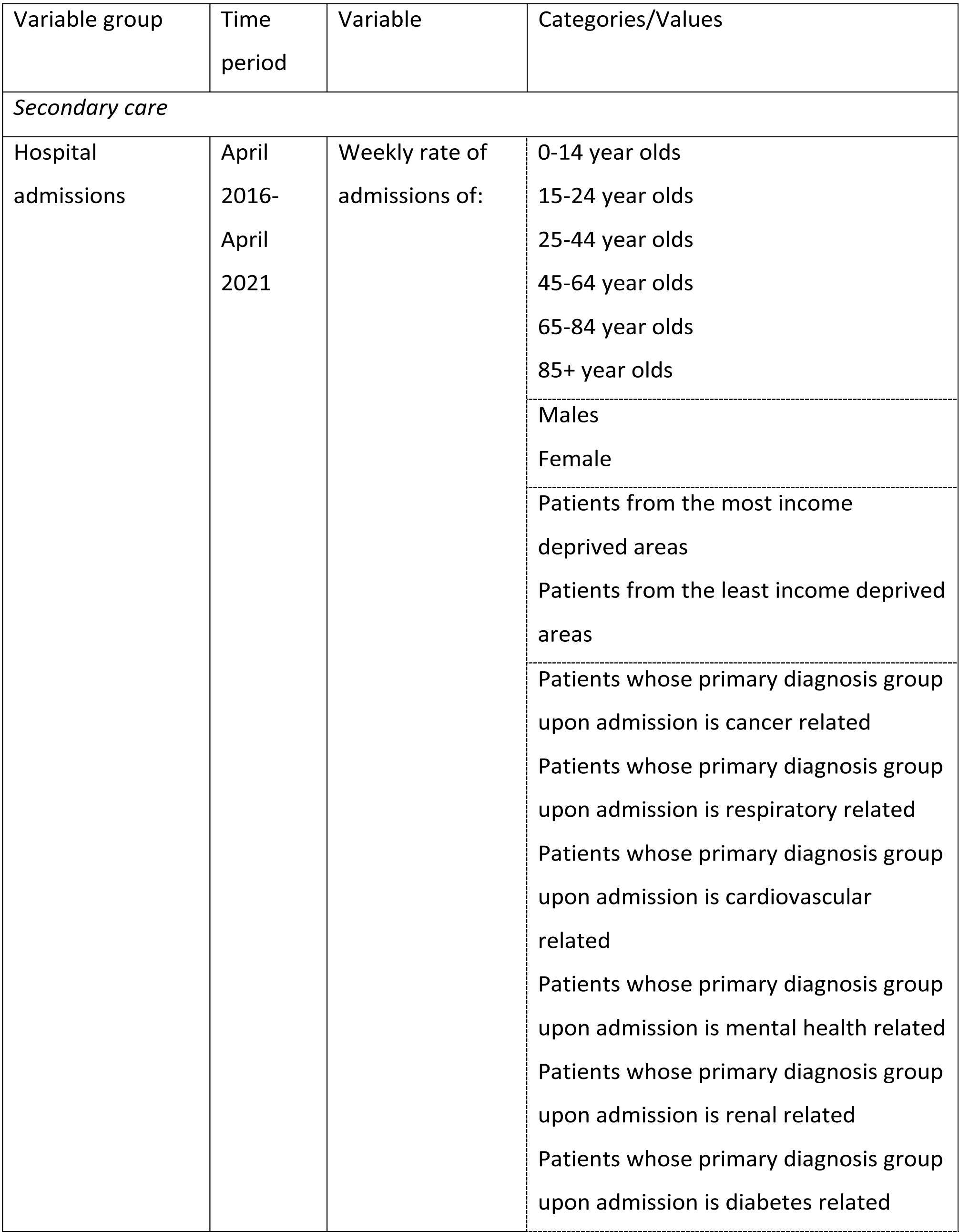
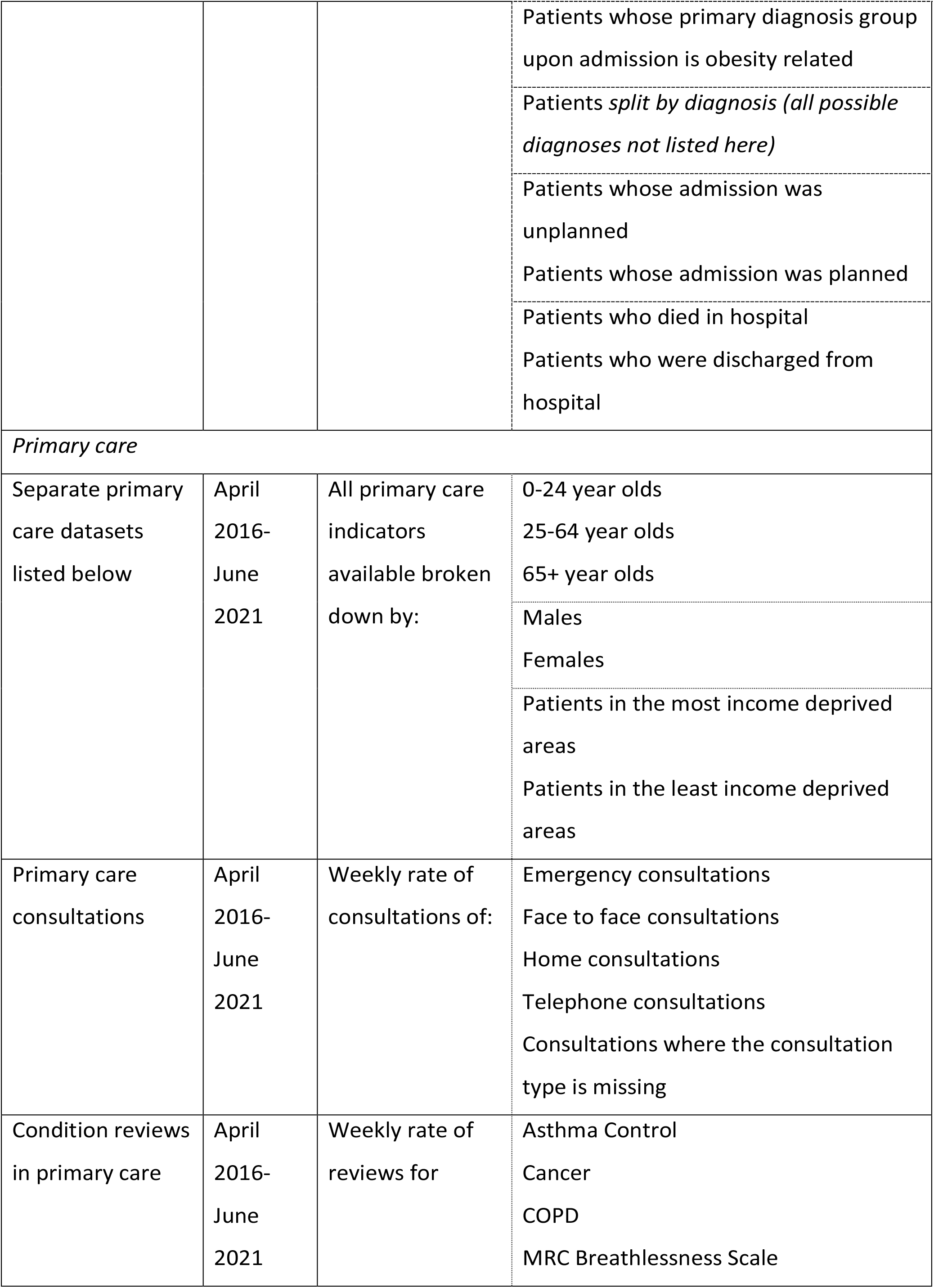
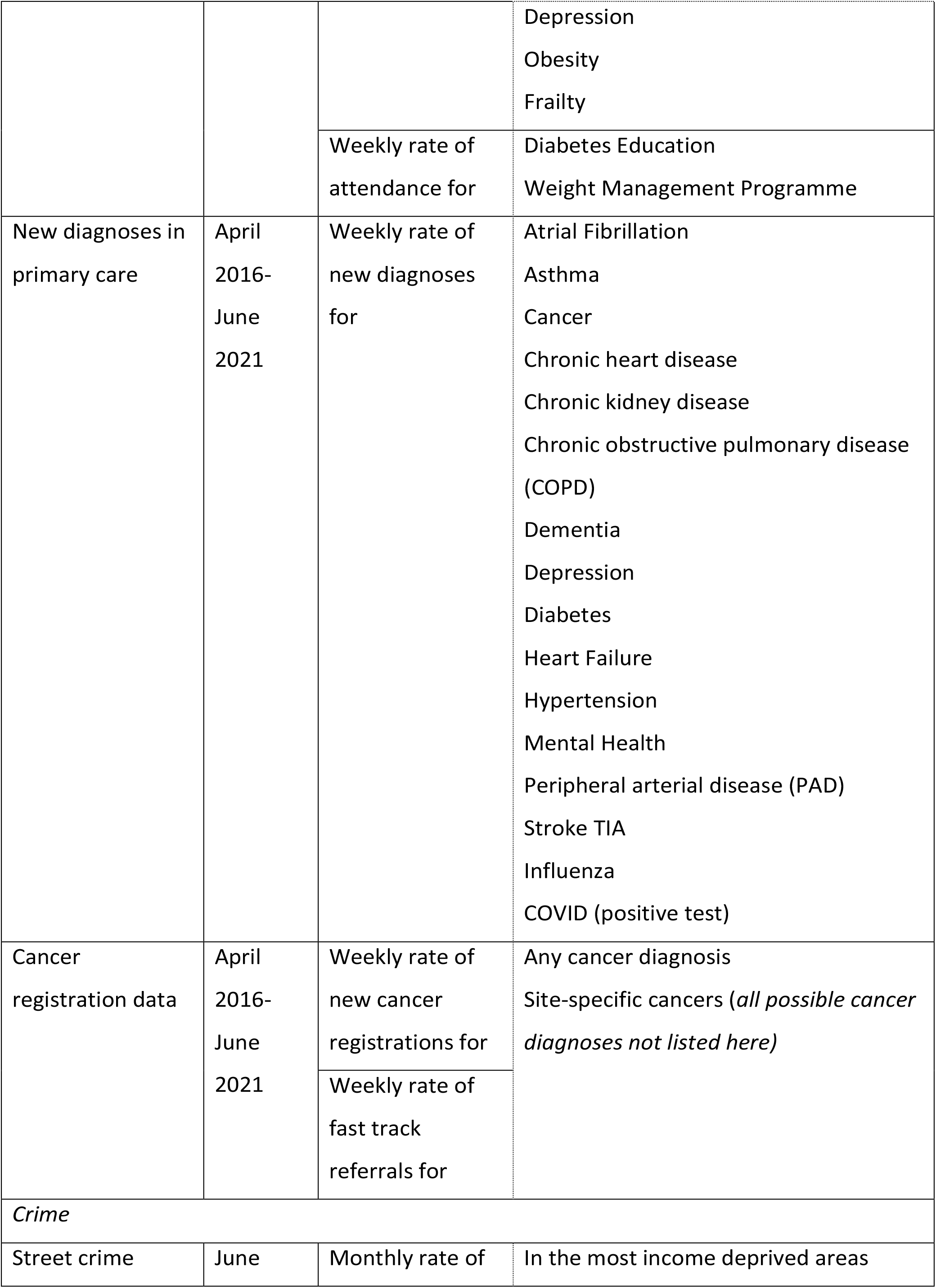
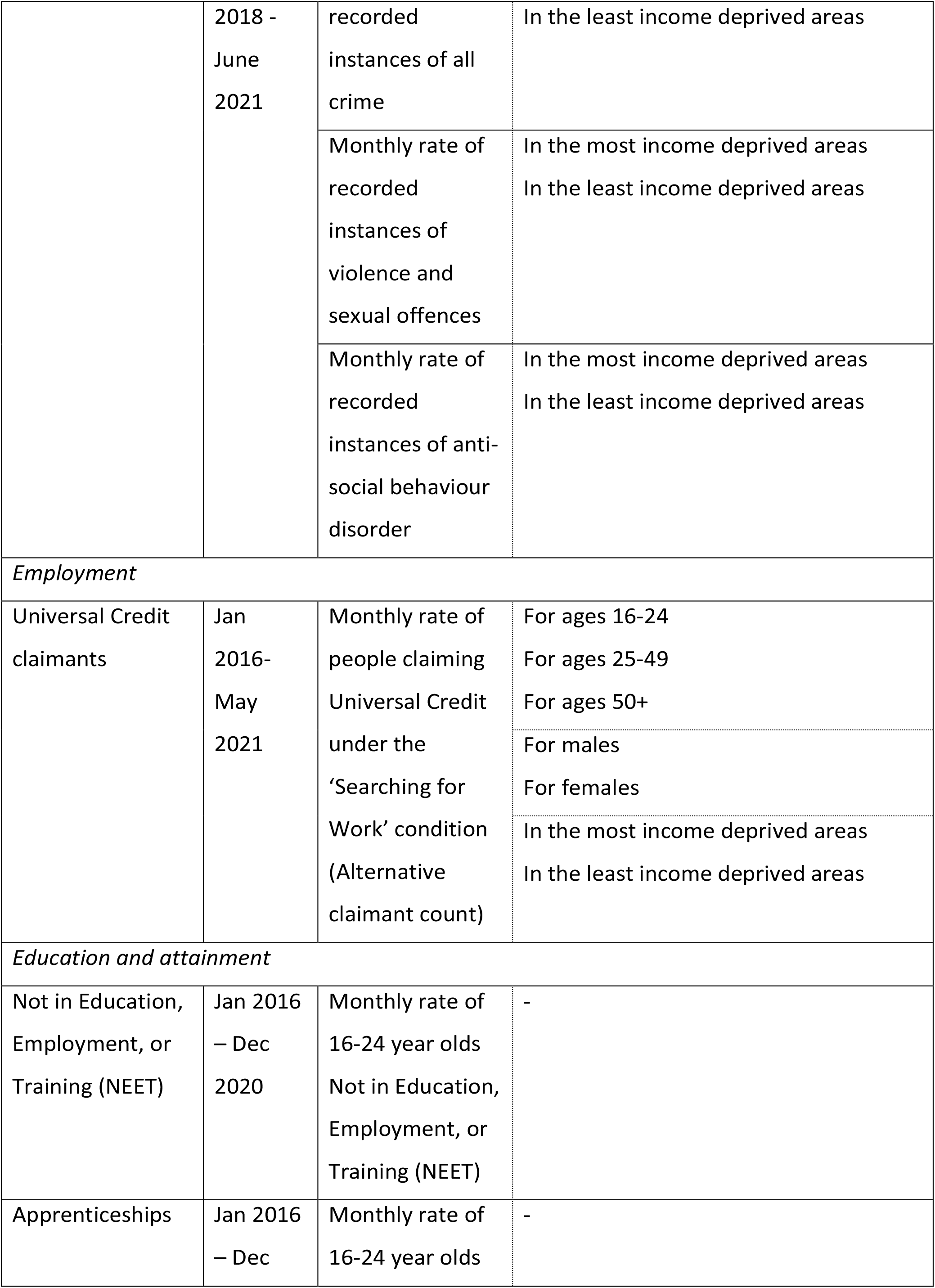
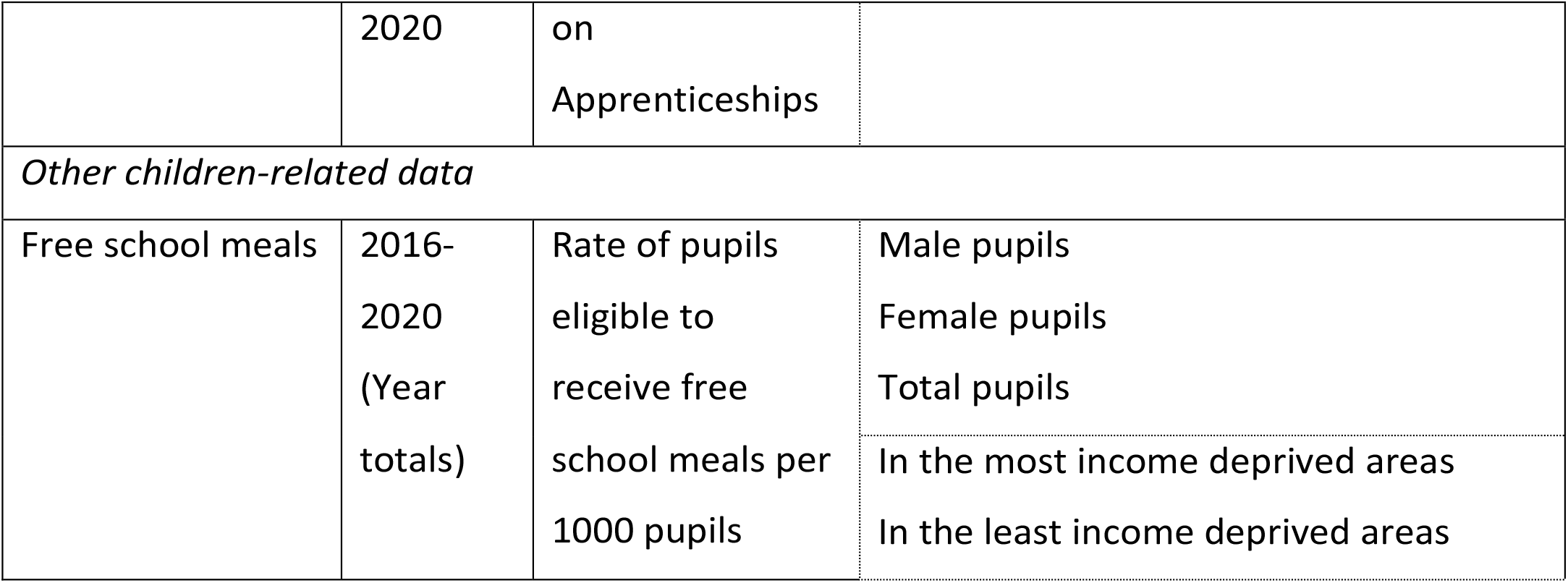
Variable descriptions

**Table 3:** Proportion of hospital admissions and primary care diagnoses in selected patient groups

| Hospital admissions |  |  |  |  |
| --- | --- | --- | --- | --- |
|  | Primary diagnosis | pre-March 2020 | post-March 2020 | p value<sup>1</sup> |
| Respiratory | No | 0.57 | 0.63 |  |
|  | Yes | 0.43 | 0.37 | <0.01 |
|  | Yes (excl. Covid) | 0.43 | 0.27 | <0.01 |
| Cardiovascular | No | 0.71 | 0.66 |  |
|  | Yes | 0.29 | 0.34 | <0.01 |
| Mental Health | No | 0.96 | 0.95 |  |
|  | Yes | 0.04 | 0.05 | 0.30 |
| Secondary Mental Health | No | 0.69 | 0.62 |  |
|  | Yes | 0.31 | 0.38 | <0.01 |
| Diagnosis in primary care |  |  |  |  |
|  | Patient group | pre-March 2020 | post-March 2020 | p value<sup>1</sup> |
| Depression | 0-24 | 0.16 | 0.22 |  |
|  | 25-64 | 0.75 | 0.67 |  |
|  | 65+ | 0.10 | 0.11 | 0.04 |
| Chronic kidney disease | Female | 0.56 | 0.30 |  |
|  | Male | 0.44 | 0.70 | <0.01 |
| Peripheral Arterial Disease | Female | 0.48 | 0.20 |  |
|  | Male | 0.52 | 0.80 | 0.02 |
| Asthma | Most Deprived | 0.66 | 0.52 |  |
|  | Least Deprived | 0.34 | 0.48 | 0.04 |
| <sup>1</sup>X<sup>2</sup> test |  |  |  |  |

#### Admissions with a primary cardiovascular diagnosis

In contrast to respiratory-related admissions, admissions primarily related to cardiovascular disease dropped sharply after March 2020 but appear to have increased again to normal levels by Autumn 2020.

#### Admissions related to mental health

Further patterns were observed for mental health-related hospital admissions. There was no apparent immediate impact in admissions of patients with a primary diagnosis of mental health following March 2020, although in Spring 2021 there are indications that admissions begin to rise again, although this increase is no more extreme than what has been experienced in the pre-COVID time (e.g. 2017).

However, there has been a sustained fall in admissions of patients with a secondary diagnosis of mental health in the post-March 2020 period. While the rate of Fleetwood population being admitted with a secondary mental health diagnosis fell, the proportion of all hospital admissions in this diagnosis category actually increased in the post-March 2020 period (31% to 38%, p<0.01) (Table 3). This likely indicates that admissions with a secondary mental health diagnosis did not fall as much as overall hospital admissions post-March 2020, or as much as other health conditions common to hospital admissions pre-March 2020 (e.g. non-COVID respiratory).

### Primary care

In primary care, there was a fall in the incidence rate of new diagnosis across several different diagnostic groups, although the patterns in the post-March 2020 period vary depending on diagnostic and demographic groups. Examples of differing patterns post-March 2020 are shown in Figure 3. These plots show the trend over time for the rate of new diagnoses in primary care for chronic obstructive pulmonary disease (COPD) and chronic kidney disease (CKD), both of which saw considerable and sustained falls in recorded new diagnoses. Diagnosis of hypertension also saw a sudden fall in Spring 2020, although the rates of new diagnoses have been increasing again since Summer 2020.

**Figure 3:**
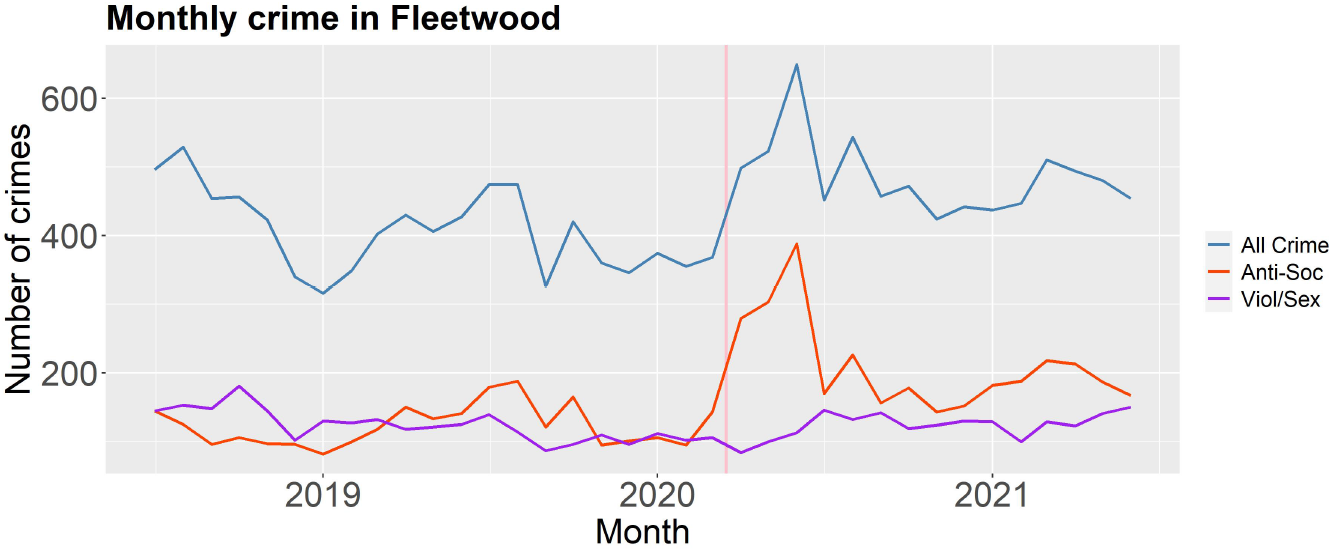
Number of monthly street crimes in Fleetwood stratified by the three largest crime categories

The changes in rates of new diagnoses between the pre- and post-March 2020 period were compared for each diagnostic group across different sexes, age groups, and areas of income deprivation (Table 3). There were significant differences in the proportion of diagnoses for depression among young people (aged up to 24 years), which has grown in the post-March 2020 period. Additionally, a lower proportion of new diagnoses of chronic kidney disease (0.30 vs. 0.56, p<0.01) and peripheral arterial disease (0.48 vs. 0.20, p=0.02) were among women in the post-March 2020, the reverse of the relationship with sex and diagnosis in the years previous (Table 3). The proportion of new diagnoses for asthma among patients living in the most income deprived areas appears to have fallen in the post-March 2020 period (0.66 vs. 0.52, p=0.04).

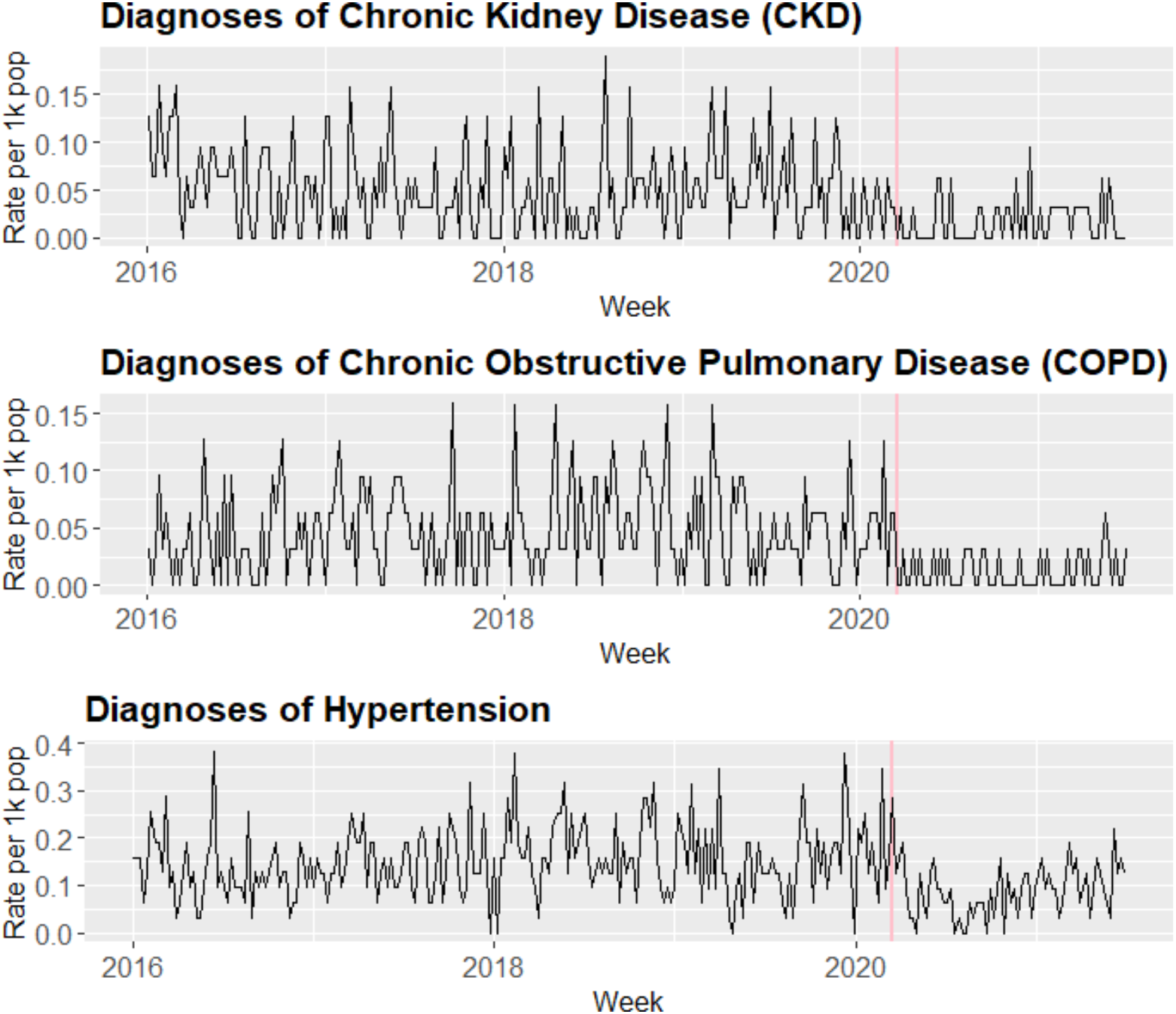

### Cancer

While there were some changes in the characteristics of patients in Fleetwood registered as having cancer, or receiving a cancer fast track referral, between the pre- and post-March 2020 temporal periods, the differences did not reach statistical significance in univariate tests.

### Crime

The number of street crime incidents in Fleetwood recorded by the police saw an increase in 2020, particularly inthe first half of that year. As the plots in Figure 4 suggest, the trend in overall street crime in 2020 appears to be related to spikes in the number of anti-social behaviour incidents in that period, which likely include the breaking of COVID-19-related restrictions.

**Figure 4:**
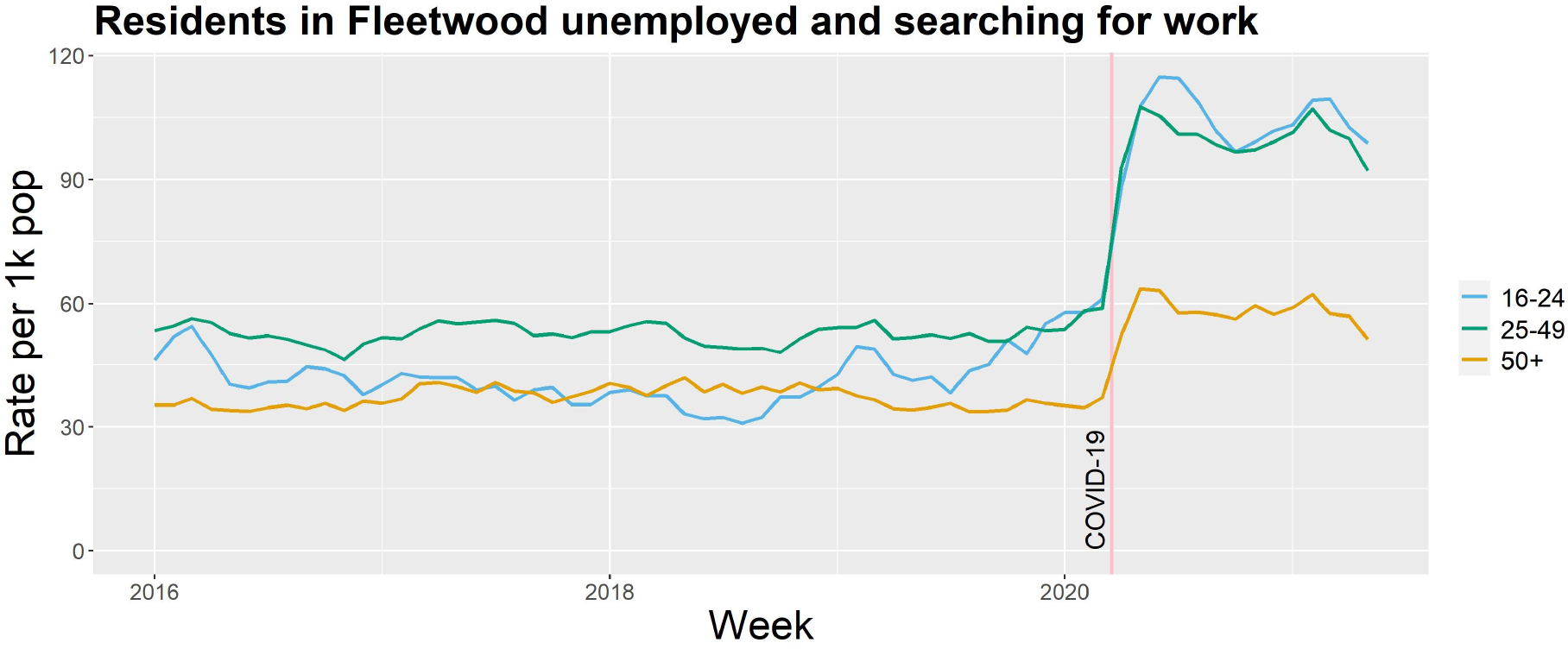
Rate of Fleetwood residents claiming Universal Credit under the ‘Searching for Work’ condition (Alternative Claimant Count)

While in the pre and post-March 2020 period, most crimes occurred in the most income deprived areas, the proportion of ‘anti-social behaviour’ and ‘violence and sexual offences’ crimes occurring in the most deprived areas fell post-March 2020, with a corresponding proportion increase in the least deprived areas (Table 4). Chi-squared tests indicated that this difference was only statistically significant for anti-social behaviour crimes (90.19% vs 88.75% in Most Deprived Areas, p<0.01) (Table 4).

**Table 4:**
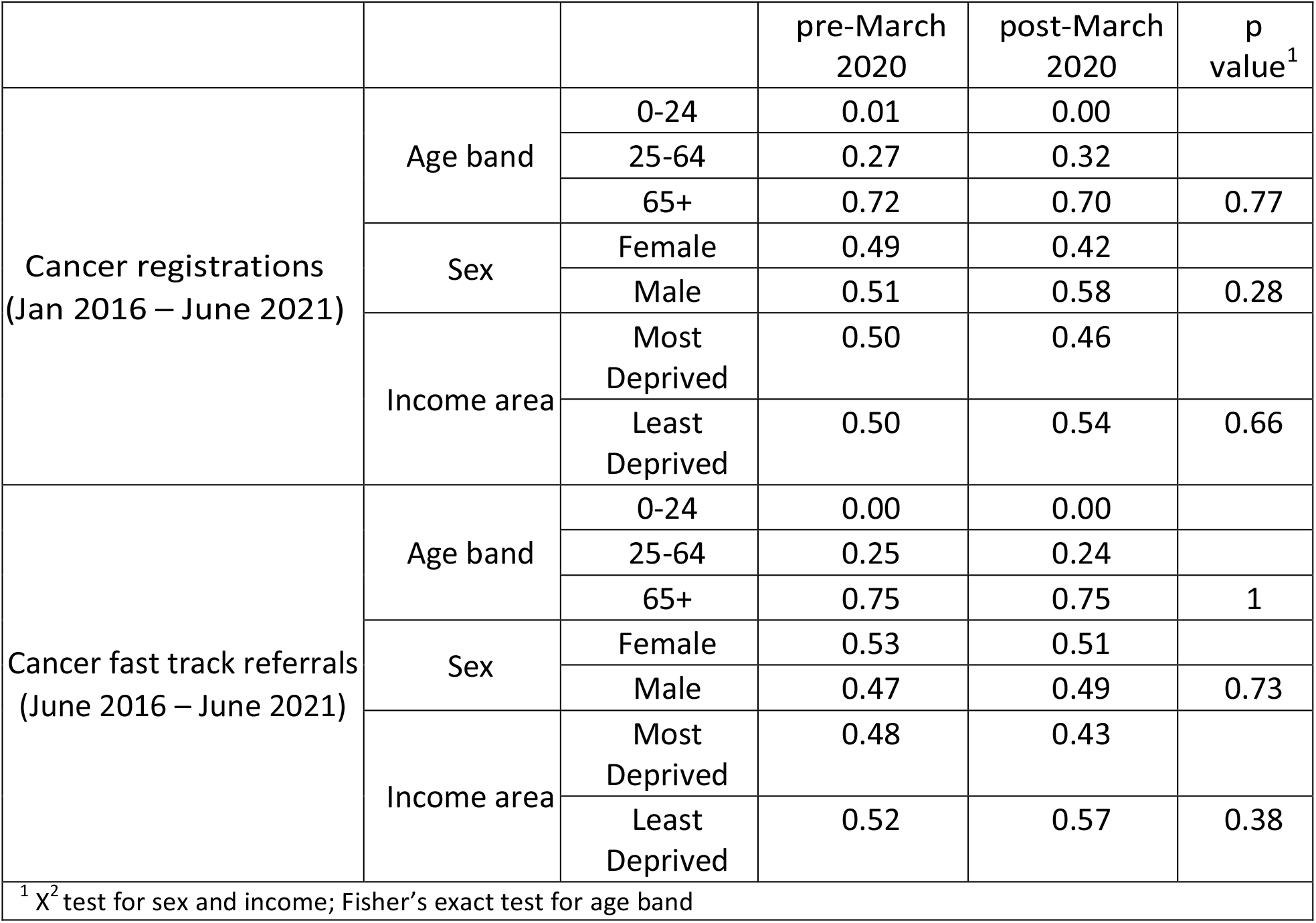
Proportion of cancer registrations and fast track referrals in patient groups

|  |  |  | pre-March<br>2020 | post-March<br>2020 | p<br>value<sup>1</sup> |
| --- | --- | --- | --- | --- | --- |
| Cancer registrations<br>(Jan 2016 – June 2021) | Age band | 0-24 | 0.01 | 0.00 |  |
|  |  | 25-64 | 0.27 | 0.32 |  |
|  |  | 65+ | 0.72 | 0.70 | 0.77 |
|  | Sex | Female | 0.49 | 0.42 |  |
|  |  | Male | 0.51 | 0.58 | 0.28 |
|  | Income area | Most Deprived | 0.50 | 0.46 |  |
|  |  | Least Deprived | 0.50 | 0.54 | 0.66 |
| Cancer fast track referrals<br>(June 2016 – June 2021) | Age band | 0-24 | 0.00 | 0.00 |  |
|  |  | 25-64 | 0.25 | 0.24 |  |
|  |  | 65+ | 0.75 | 0.75 | 1 |
|  | Sex | Female | 0.53 | 0.51 |  |
|  |  | Male | 0.47 | 0.49 | 0.73 |
|  | Income area | Most Deprived | 0.48 | 0.43 |  |
|  |  | Least Deprived | 0.52 | 0.57 | 0.38 |
| <sup>1</sup> X<sup>2</sup> test for sex and income; Fisher's exact test for age band |  |  |  |  |  |

### Unemployment

The rate of people receiving Universal Credit each month is comparable over time using the Alternative Claimant Count (ACC). This is an indication of the number of people who are receiving Universal Credit under the ‘searching for work’ condition, meaning they are required to look for work while receiving financial support.

Table 4 shows the monthly average ACC rate for Fleetwood in the pre- and post-March 2020 periods. It indicates there was a statistically significant increase in the rate of claimants per 1000 population post-March 2020 (27.4 vs 50.6, p*<*0.01). In the post-March 2020 period, existing differences between male and female claimants became larger, with a greater proportion of male claimants post-March 2020 (Table 4).

Where available, variables have been stratified by income deprivation area, capturing differences between areas in Fleetwood among the 20% most deprived areas nationally (‘Most’ in Table 4) and those that are less deprived comparatively (‘Least’ in Table 4). Post-March 2020, the proportion of younger claimants and those living in the least deprived areas of Fleetwood grew (Table 4). Figure 5 shows the time trends in claimant rates split by age.

**Figure 5:**
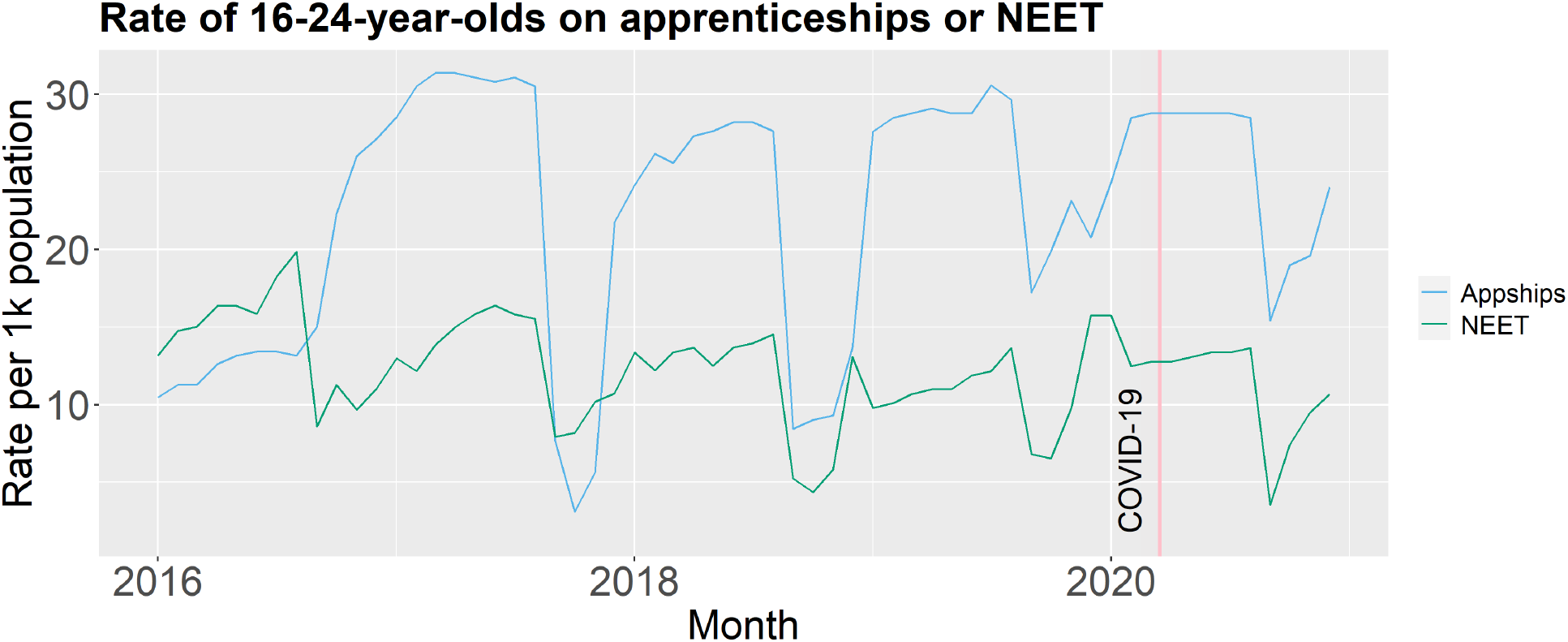
Rate of 16-24 year olds in Fleetwood on apprenticeships or NEET

### School leaver opportunities

The study dataset includes information about the rate of 16-24-year-olds in Fleetwood either enrolled in an apprenticeship or ‘Not in Education, Employment, or Training’ (NEET) from 2016 until 2020. Since 2016 the rates of young people who are NEET appear to have been decreasing and the rates on apprenticeships appears moderately stable (Figure 6). Both appear to have seasonal trends, which likely correspond to school leaving dates. The mean monthly rates for each year in Table 5 suggest that 2020 rates appear to counter the trend somewhat, with rates of NEET increasing slightly and those on apprenticeships decreasing on the 2019 figure, although these changes are very small. Furthermore, when comparing the pre and post-March 2020 period, the post-March 2020 rates are higher overall, although this is not statistically significant (Table 4).

**Table 5:**
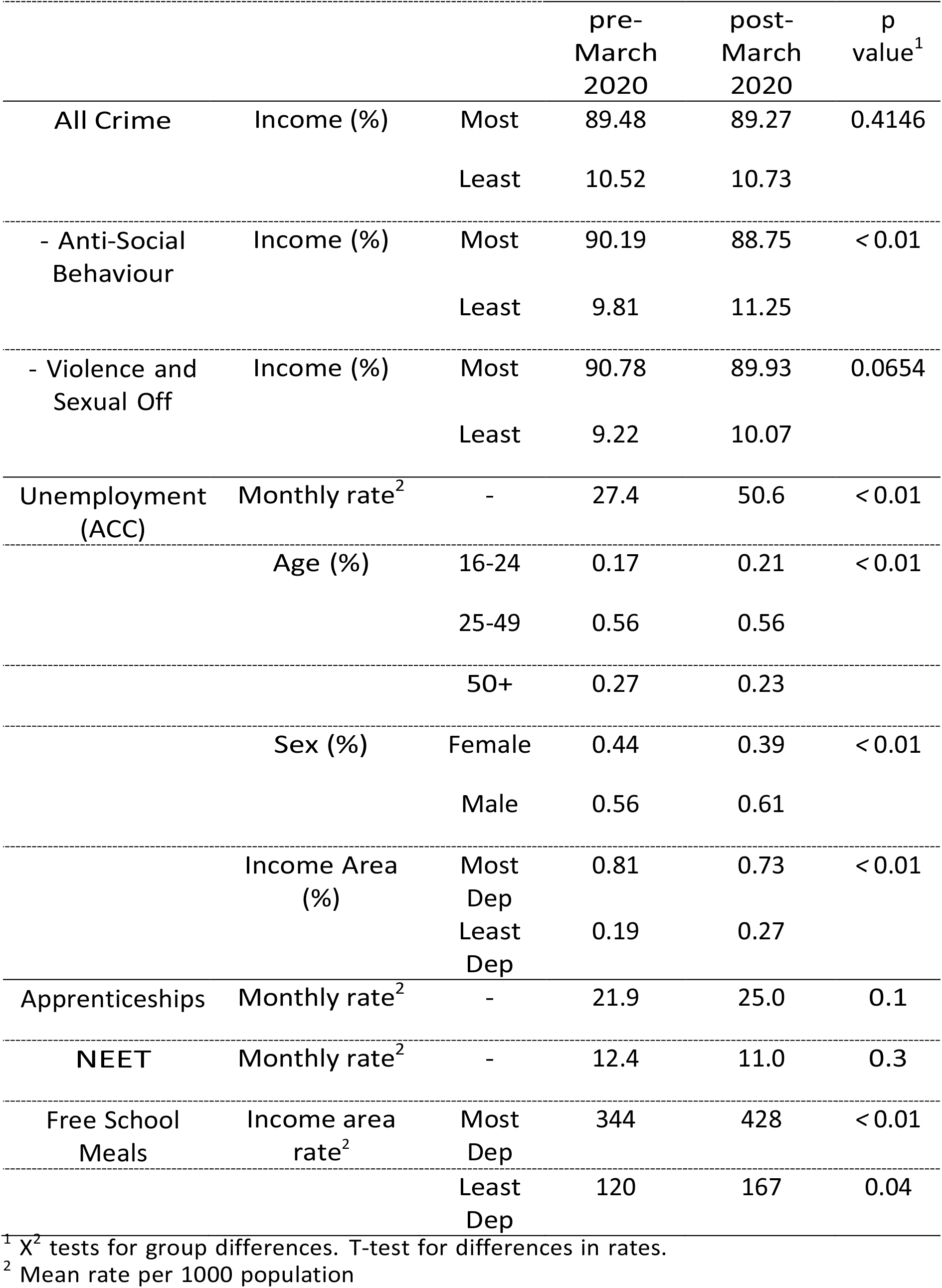
Group differences for selected social and economic indicators pre and post-March 2020

|  |  |  | pre-March<br>2020 | post-March<br>2020 | p<br>value<sup>1</sup> |
| --- | --- | --- | --- | --- | --- |
| All Crime | Income (%) | Most | 89.48 | 89.27 | 0.4146 |
|  |  | Least | 10.52 | 10.73 |  |
| - Anti-Social<br>Behaviour | Income (%) | Most | 90.19 | 88.75 | < 0.01 |
|  |  | Least | 9.81 | 11.25 |  |
| - Violence and<br>Sexual Off | Income (%) | Most | 90.78 | 89.93 | 0.0654 |
|  |  | Least | 9.22 | 10.07 |  |
| Unemployment<br>(ACC) | Monthly rate<sup>2</sup> | - | 27.4 | 50.6 | < 0.01 |
|  | Age (%) | 16-24 | 0.17 | 0.21 | < 0.01 |
|  |  | 25-49 | 0.56 | 0.56 |  |
|  |  | 50+ | 0.27 | 0.23 |  |
|  | Sex (%) | Female | 0.44 | 0.39 | < 0.01 |
|  |  | Male | 0.56 | 0.61 |  |
|  | Income Area<br>(%) | Most<br>Dep | 0.81 | 0.73 | < 0.01 |
|  |  | Least<br>Dep | 0.19 | 0.27 |  |
| Apprenticeships | Monthly rate<sup>2</sup> | - | 21.9 | 25.0 | 0.1 |
| NEET | Monthly rate<sup>2</sup> | - | 12.4 | 11.0 | 0.3 |
| Free School<br>Meals | Income area<br>rate<sup>2</sup> | Most<br>Dep | 344 | 428 | < 0.01 |
|  |  | Least<br>Dep | 120 | 167 | 0.04 |

<sup>1</sup> X<sup>2</sup> tests for group differences. T-test for differences in rates.<sup>2</sup> Mean rate per 1000 population

**Table 6:** Mean rate (per 1k) of 16-24 year olds on apprenticeships or NEET

| Year | Apprenticeships | NEET |
| --- | --- | --- |
| 2016 | 15.8 | 14.2 |
| 2017 | 23.6 | 12.9 |
| 2018 | 21.3 | 11.3 |
| 2019 | 26.1 | 10.8 |
| 2020 | 25.3 | 11.5 |

**Figure 6:**
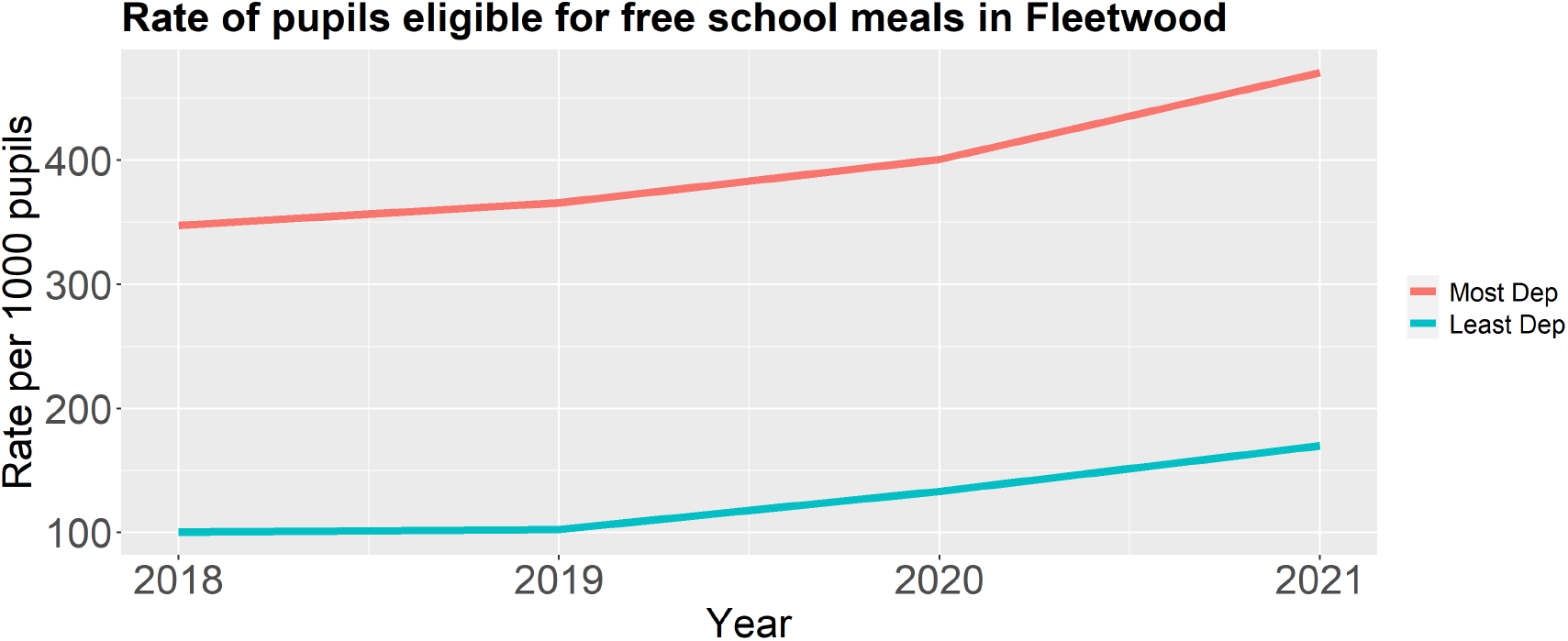
Rate of school pupils eligible to receive free school meals in Fleetwood by income deprivation area

### Free School Meals

As the plot in Figure 7 and summary statistics in Table 4 show, the mean rate of pupils entitled to receive free school meals per 1000 pupils grew in the post-COVID-19 period in both the most and least income deprived areas in Fleetwood. While the numbers of pupils eligible for free school meals is much higher in the most deprived areas both pre- and post-COVID-19, the rate of pupils eligible for free school meals per 1000 pupils increased by 40% in the least deprived areas (120 pre vs 167 post, p=0.04), compared to 24% in the most deprived areas (344 pre vs 428 post, p<0.01).

## Discussion

The findings reported in this manuscript are those from an exploratory analysis of routinely collected secondary health and socioeconomic data for Fleetwood residents. Temporal and spatial data were collected to examine differences in outcomes in different areas of Fleetwood, a socially deprived coastal town in North West England. While exploring mostly univariate associations between key variables of interest, these initial findings indicate the degree to which the health and social wellbeing of residents of Fleetwood have been impacted in the first 18 months following the COVID-19 pandemic. The results point towards some key areas that call for more comprehensive analysis using survival, multivariate time series and spatial analyses. These areas of interest include the large and sustained falls in hospital admissions for respiratory-related conditions, the changes in the age and sex demographics of incidence of diagnosis in primary care, and indications of changes to some social outcomes in some of the least deprived areas of the towns.

The time trends of hospital admissions and primary care diagnoses, along with the statistical test of pre- and post-March 2020 differences, indicate the varied nature of the pandemic’s impact on different patient populations. This corresponds to other study findings, suggestive of condition-varying patterns in healthcare use in the first six months of 2020 (4). Covering a longer time period, the data in this ongoing study indicates longer-term trends (18 months) of healthcare use in the post-March 2020 period. Some of the most striking findings are the sustained falls in some healthcare outcomes related to non-COVID-19 respiratory conditions in this period. By June 2021, there were still no signs of the rate of new chronic obstructive pulmonary disease (COPD) diagnoses increasing to pre-March 2020 levels. However, low rates likely reflect the barrier to confirming a diagnosis of COPD using spirometry, a breathing examination temporarily not in use because of COVID-19 infection concerns (30). The Association for Respiratory Technology and Physiology (ARTP) and the Primary Care Respiratory Society (PCRS) estimate the number of patients awaiting a spirometry diagnosis to be around 200–250 patients per 500,000 population (30). While these will not all be COPD diagnoses, many patients with suspected COPD are likely to have been put on a ‘waiting list’ to be formally diagnosed before being placed on a COPD register. While some falls in diagnosis may relate to system barriers rather than patients not being seen in primary care, they could nonetheless lead to delays in care management and quicker deterioration for some patients. Additionally, the rate of hospital admissions where the primary diagnosis is a non-COVID-19 respiratory condition has also remained consistently low throughout the study period, suggesting that falls in admissions documented early on in the pandemic are persisting into 2021 (7).

This exploratory analysis also points towards changing patterns to patient and population demographics of some health and socioeconomic outcomes in the pre- and post-March 2020 periods. Some of the associations in the pre-March 2020 period have been reversed, as is the case for women making up a much lower proportion of diagnoses for chronic kidney disease or peripheral arterial disease, a pattern not easily explained. Other trends may have more apparent explanations. For example, the increase in the proportion of depression diagnoses among young people aligns to a growing research literature pointing towards the impact the pandemic has had on that population group’s mental wellbeing, who have experience substantial disruptions to school, social, and working life (31,7). Within this population, sub-groups may be particularly affected. There is evidence to suggest that among 16-25 years old, being female and having low social support were both predictors of mental distress during the pandemic (32). However, given that some of the most significant contributors to mental distress among young people during COVID have been reduced social support, activities, and increased loneliness, it may be that mental wellbeing will increase with greater social interactions (32). Assessing the longevity of this impact will require longitudinal research that follows this population over many years.

Several socioeconomic outcomes, including the rate of children eligible to receive free school meals, the numberof street crimes, and unemployment, appear to have increased in the medium term in areas classified as the’least deprived’ in Fleetwood. This does not necessarily contradict suggestions made elsewhere that the COVID-19 pandemic will exacerbate social inequalities in health and social outcomes, with a greater impact on those experiencing socioeconomic disadvantage (19). The apparent changes in the ‘least deprived’ areas could indicate the pre-pandemic vulnerability of population groups experiencing moderate but not severe financial insecurity. When compared to all other neighbourhoods in England, the ‘least deprived’ areas in Fleetwood sit in the middle of national deprivation rankings. Other studies have suggested that those in middle-income housing, particularly those who were in work but on low pay, may have experienced the greatest financial disruption to their lives (17). It is possible that some residents in the ‘least deprived’ areas of Fleetwood were more vulnerable than others to the economic impacts of the pandemic.

## Conclusion

There are early indications that the COVID-19 pandemic has had an impact on several health and social outcomes in Fleetwood, a predominantly socially deprived town in North West England. An exploratory analysis of these impacts suggests they may be patterned differently depending on health conditions, patient and population demographics, and area income deprivation. Examining these trends in multivariate analyses will test these associations and establish the strength of the impact on residents in this coastal community.

## Data Availability

Access to data produced in the present study can be requested from the authors. Access will only be granted subject to approval by authors and data providers.

